# Genetic study of circulating cytokines offers insight into the determinants, cascades and effects of systemic inflammation

**DOI:** 10.1101/2020.10.26.20219477

**Authors:** Ville Karhunen, Dipender Gill, Rainer Malik, Mark J. Ponsford, Ari Ahola-Olli, Areti Papadopoulou, Saranya Palaniswamy, Shivaprakash Jagalur Mutt, Sylvain Sebert, Minna Männikkö, Juha Auvinen, Juha Veijola, Karl-Heinz Herzig, Markku Timonen, Sirkka Keinänen-Kiukaanniemi, Martin Dichgans, Marko Salmi, Sirpa Jalkanen, Terho Lehtimäki, Veikko Salomaa, Olli Raitakari, Simon A Jones, Konstantinos K. Tsilidis, Marjo-Riitta Järvelin, Abbas Dehghan

## Abstract

Cytokines are the signalling molecules that underlie inflammatory processes. Here, we performed genome-wide association study (GWAS) analyses of 47 circulating cytokines in up to 13,365 individuals to identify protein quantitative trait loci (pQTL). Applying a novel approach, we incorporated pQTL and expression quantitative trait loci (eQTL) data of 10,361 tissue samples in 635 individuals to identify biologically plausible genetic instruments to proxy the effect of cytokines. Using Mendelian randomization analysis, we explored the causal determinants of inflammatory cytokines, investigated inflammatory cascades and evaluated their effects on 20 diseases. We show evidence of body mass index (BMI), smoking and systolic blood pressure (SBP) being associated with inflammation, and specifically BMI affecting levels of active PAI-1, HGF, MCP1, sE-Selectin, sICAM1, TRAIL, IL6 and CRP. Our analysis highlights a key role of VEGF in influencing the levels of eight other inflammatory cytokines. Finally, we report evidence of sICAM affecting waist circumference and risk of major depressive disorder, evidence for TRAIL affecting the risk of cardiovascular diseases, breast and prostate cancer, and evidence for MIG affecting the risk of stroke. Overall, our results offer insight into inflammatory mediators of BMI, smoking and SBP, pleiotropic effects of VEGF, and circulating cytokines that increase the risk of cancer, cardiovascular, metabolic and neuropsychiatric diseases. All the studied cytokines represent pharmacological targets and therefore offer opportunities for clinical translation in diseases with inflammatory components.

## Introduction

Cytokines, chemokines, growth factors and interferons (hereafter cytokines) are circulating protein signalling molecules that underlie inflammatory processes(1). Their levels are influenced by metabolic traits, and in turn have implications for various diseases(2). Cytokines are already targeted clinically for the prevention and treatment of a range of autoimmune and inflammatory processes(3), with further promise for cardiovascular(4), cancer(5), and neuropsychiatric diseases(6). Elucidation of the mechanisms relating cytokines to disease risk is important for the prioritisation of agents for study in clinical trials.

Availability of genotyped populations in which circulating cytokines have been measured has allowed for an investigation into the genetic determinants of systemic cytokine levels(7, 8). Similarly, variants associated with expression of cytokine genes across different tissues have been described(9). These genetic variants that associate with circulating cytokine levels and gene expression may be used as instrumental variables in the Mendelian randomization (MR) paradigm to study potential causal effects of inflammatory cytokines on disease risk(10, 11). In MR, randomly allocated genetic variants are used as unconfounded proxies for the effects of the exposure. These genetically predicted exposures are then evaluated for their association with the outcome of interest. MR can help overcome the confounding and reverse causation that hamper causal inference in conventional observational research(10, 12), and adds to the triangulation of causal evidence from multiple sources. With an estimated median cost of $985 million required to take a new medicine to market(13), genetic data offers an opportunity to prioritise target candidates while saving on costs(14).

In this study, we conducted the largest genome-wide association studies (GWAS) of 47 circulating cytokines (Table 1) in up to 13,365 individuals and performed MR analyses to investigate the effect of cardiometabolic traits on their levels. Further, incorporating gene expression GWAS data from 10,361 samples on 53 tissues in 635 individuals, we aimed to identify genetic instruments with *a priori* high biological relevance to proxy the effect of varying circulating cytokine levels. We applied these instruments to explore inflammatory cascades and effects on cancer, cardiovascular, metabolic and neuropsychiatric outcomes.

**Table 1.**
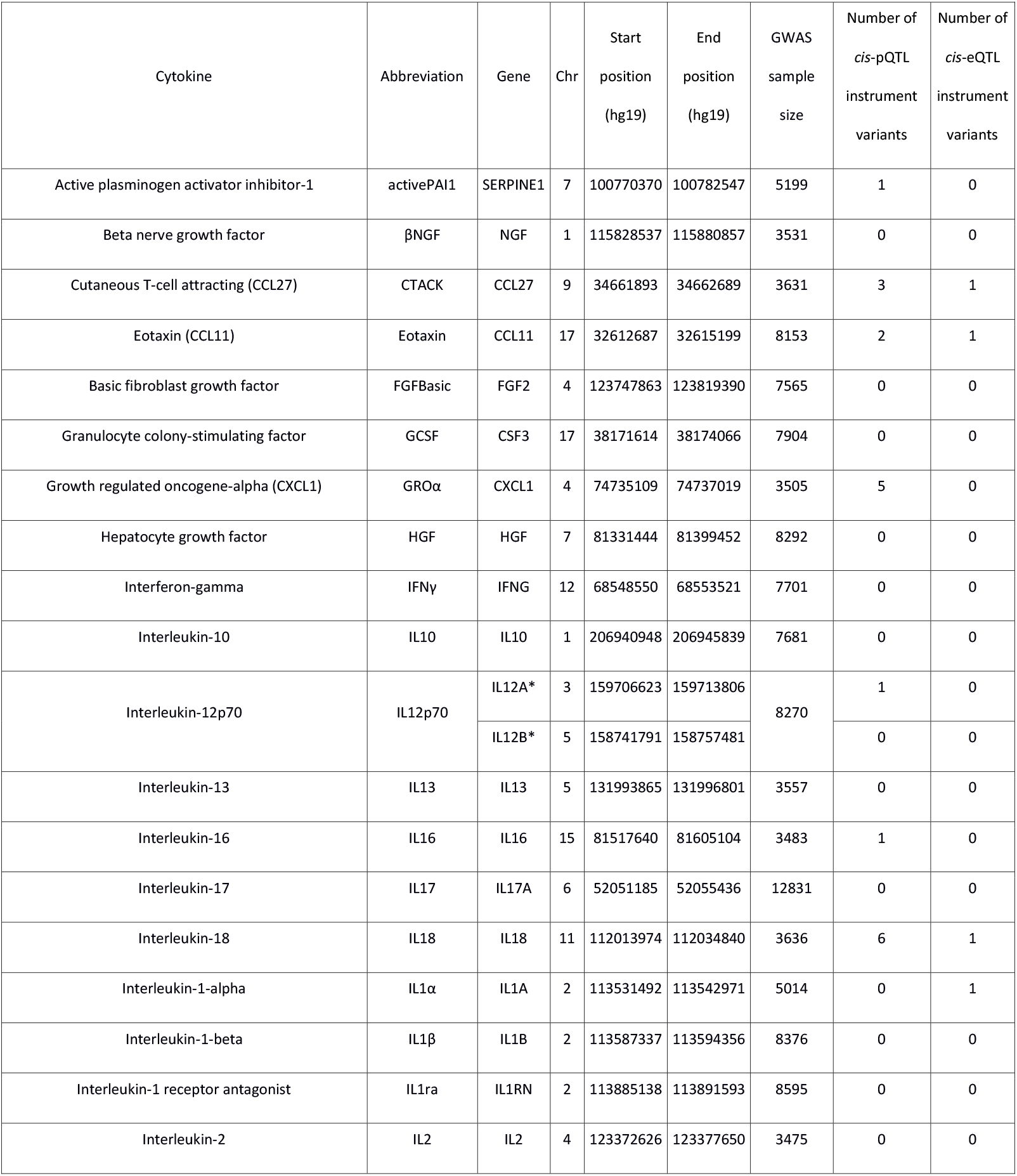
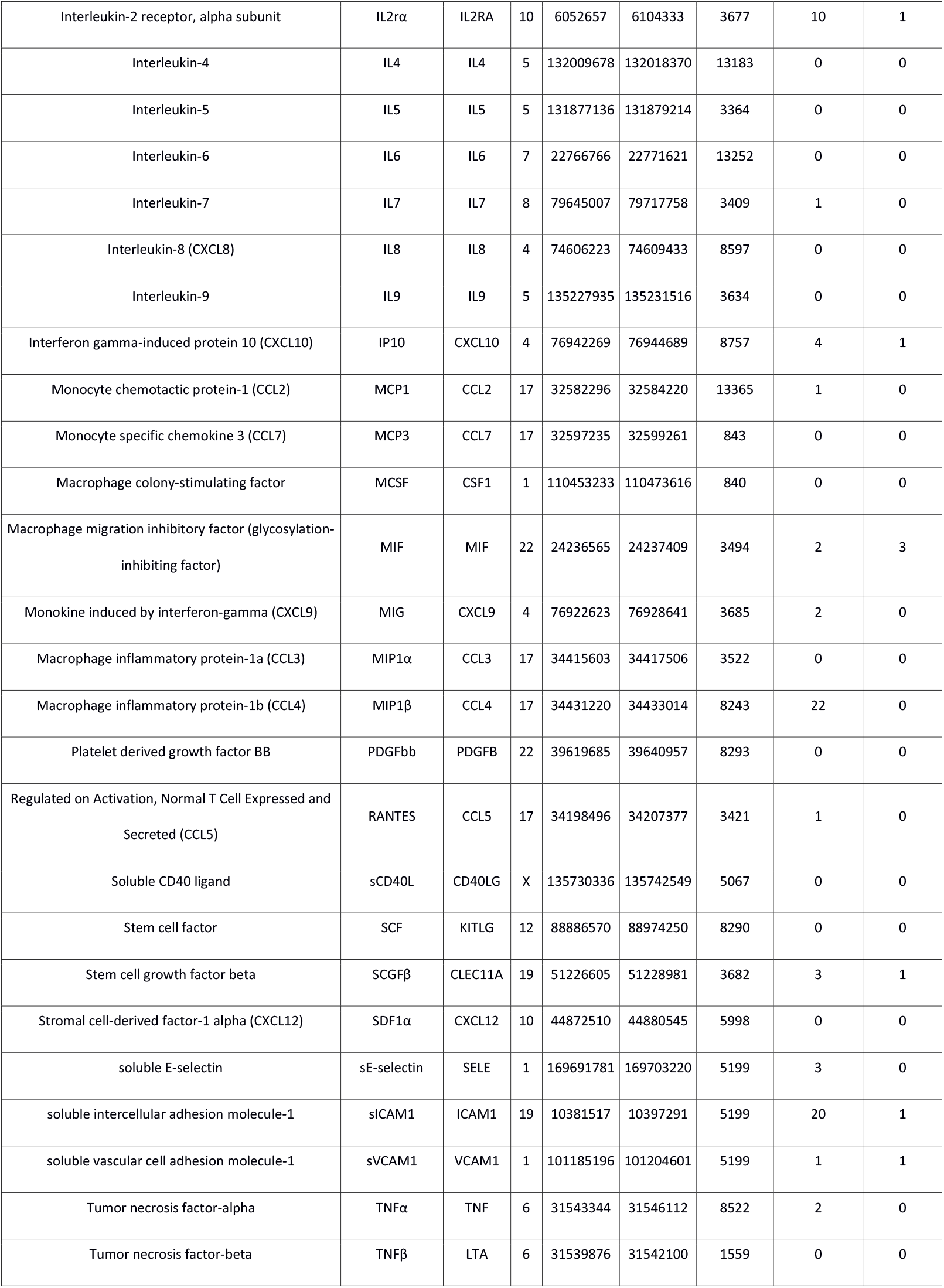
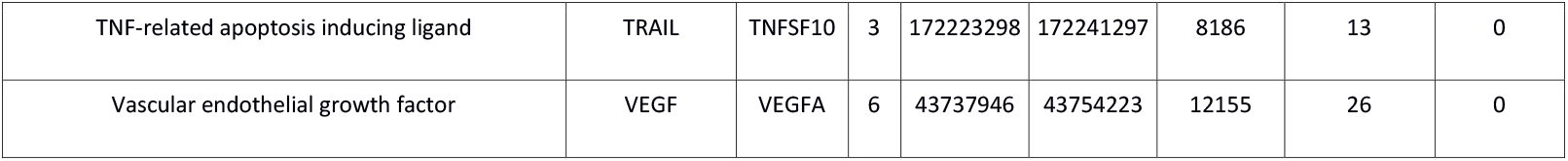
Cytokines examined in this study. Chr: chromosome; GWAS: genome-wide association study; pQTL: protein quantitative trait loci; eQTL: expression quantitative trait loci. *Interleukin-12p70 has two coding genes.

## Results

Figure 1 shows the study design. We conducted GWAS on the circulating levels of 47 cytokines (Manhattan and QQ-plots are presented in Supplementary Figures 1-47) and used the summary statistics for the subsequent MR analysis. First, we examined potential causal effects of cardiometabolic traits on circulating cytokine levels. We found positive associations (*P* < 0.0011, applying a Bonferroni correction for testing of multiple cytokines) between:

i. genetically predicted body-mass index (BMI) and circulating levels of C-reactive protein (CRP), hepatic growth factor (HGF), interleukin (IL) 16, tumour necrosis factor related apoptosis-inducing ligand (TRAIL), monocyte chemoattractant protein-1 (MCP1), active plasminogen activator inhibitor-1 (activePAI1), soluble E-selectin (sE-selectin) and soluble vascular cell adhesion molecule-1 (sICAM1),
ii. genetically predicted smoking and levels of CRP and sICAM1, and
iii. genetically predicted systolic blood pressure (SBP) and CRP (Figure 2, Supplementary Figure 48, Supplementary Table 1).

**Figure 1.**
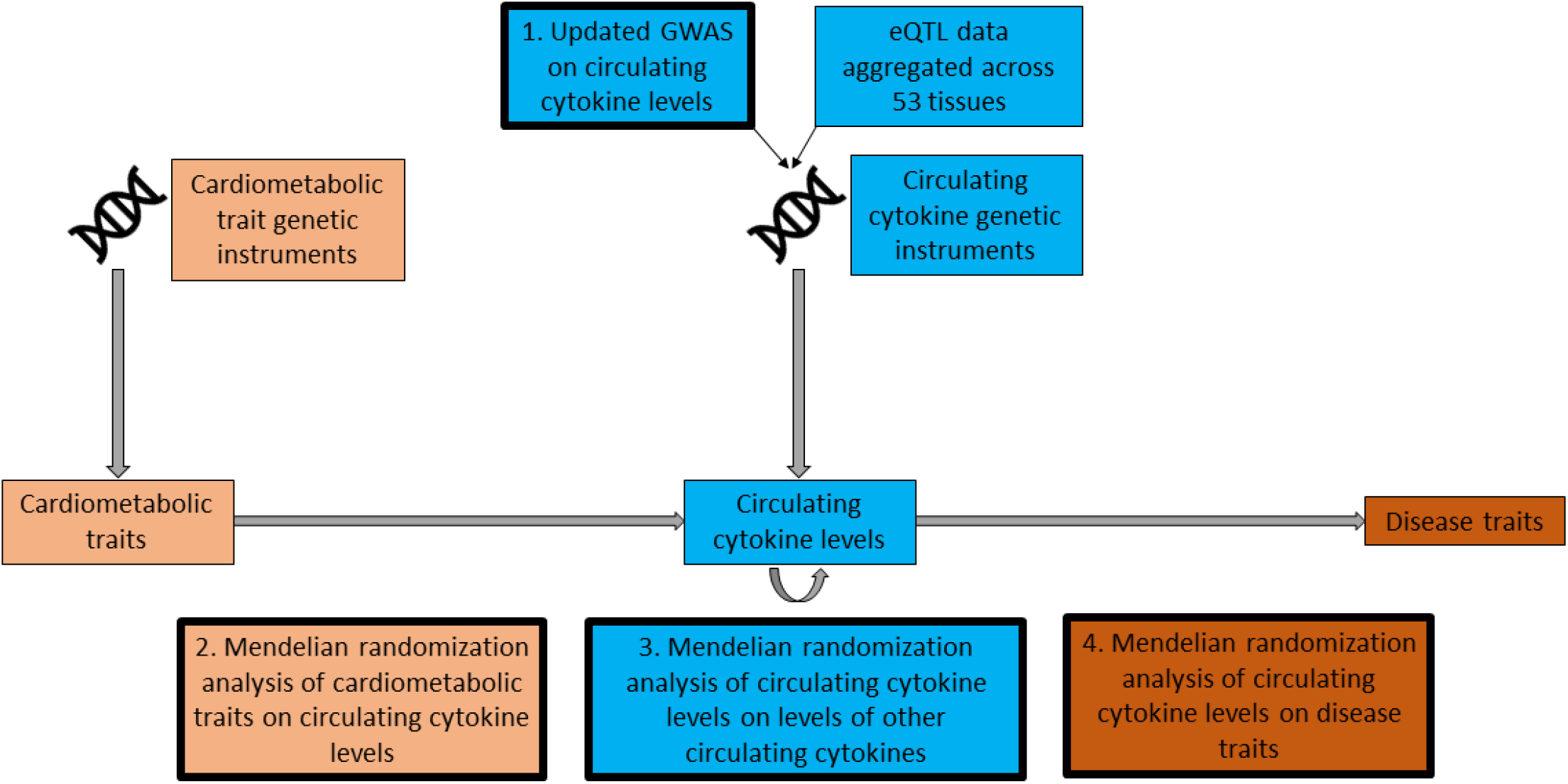
Schematic presentation of the study. The analyses conducted within this study are highlighted with bolded box edges.

**Figure 2.**
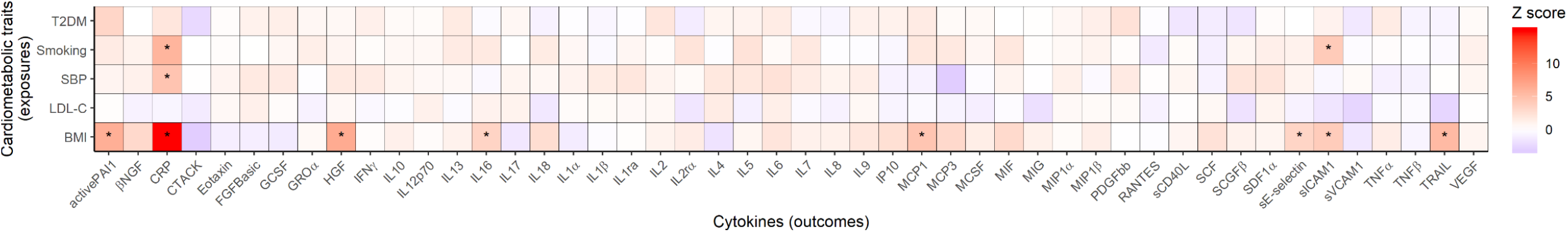
Mendelian Randomization effect size estimates (Z-scores) of genetically predicted cardiometabolic traits on circulating cytokine levels. The asterisks represent significance after Bonferroni correction for testing of multiple cytokines (*P* < 0.0011 (0.05/number of cytokines)). BMI: body-mass index; LDL-C: low-density lipoprotein cholesterol; SBP: systolic blood pressure; T2DM: type 2 diabetes mellitus. CRP: C-reactive protein. The abbreviations for other cytokines are given in Table 1.

To examine the impact of circulating cytokine levels on each other and on disease outcomes, we selected genetic instruments for circulating cytokine levels using two different criteria. In the first approach, we selected instrument variants within 500 kb of their corresponding gene locus and associated with their circulating levels at *P* < 1×10^−4^, which we term *cis*-protein quantitative trait loci (*cis*-pQTL) variants (Table 1). In the second approach, we chose instrument variants within 500 kb of the corresponding gene locus and associated with both its gene expression aggregated across tissues at *P* < 1×10^−4^, and its circulating cytokine levels at *P* < 0.05, which we term *cis*-expression quantitative trait loci (*cis*-eQTL) variants (Table 1). The *cis*-pQTL and *cis*-eQTL instrument variants were available for 22 and 10 cytokines, respectively, with both types of instruments available for nine cytokines (Table 1). F-statistics (as a measurement of instrument strength, based on circulating protein levels) for all instrument variants ranged from 15 to 928 for *cis*-pQTLs (Supplementary Table 2) and from 5 to 178 for *cis*-eQTLs (Supplementary Table 3).

Next, we investigated the potential causal effects of circulating cytokine levels on each other. There was MR evidence (*P* < 0.0011) for associations between higher genetically predicted circulating cytokine levels and both increased and decreased levels of other circulating cytokines when using the *cis*-pQTL or *cis*-eQTL instrument selection criteria (Figure 3, Supplementary Figure 49, Supplementary Table 4). Most associations were seen for genetically predicted vascular endothelial growth factor (VEGF) and IL18, which were associated with circulating levels of eight and four other cytokines, respectively (Figure 3).

**Figure 3.**
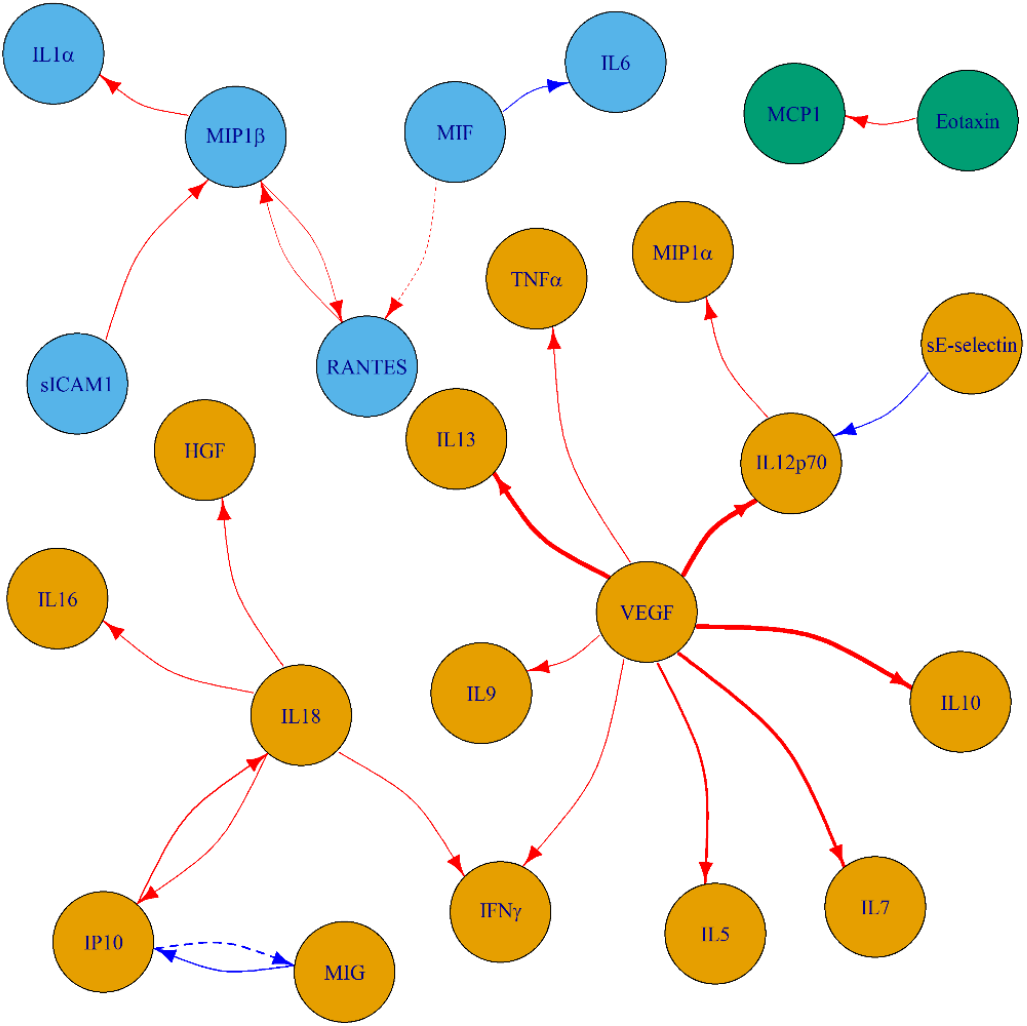
Mendelian randomization results of genetically predicted cytokine levels on levels of other circulating cytokines when considering *cis*-protein quantitative trait loci (solid lines) and *cis*-expression quantitative trait loci (dashed lines) instruments. The results are plotted only for effects with *P* < 0.0011 (0.05/number of cytokines). Red and blue lines indicate positive and negative associations respectively. The thickness of the line represents the absolute value of the effect size. The colours of the cytokines represent separate cytokine groups based on the results.

We used the GWAS-pairwise colocalization method(15) to further investigate the putative causal effects of circulating cytokine levels on each other based on our identified MR associations. We investigated proportionality of associations at the exposure gene locus with both the exposure and outcome traits, and thus any evidence for colocalization would further support a causal relationship. The results showed evidence of colocalization (posterior probability for shared variant > 0.9) for the associations of VEGF with interferon gamma (IFNγ), IL10, IL12p70, IL13, IL5 and IL17 at the *VEGF* gene locus, and for the association between monokine induced by interferon-gamma (MIG) and interferon gamma-induced protein 10 (IP10) at the *MIG* gene locus (Figure 4), providing supporting evidence for causality between the corresponding cytokines.

**Figure 4.**
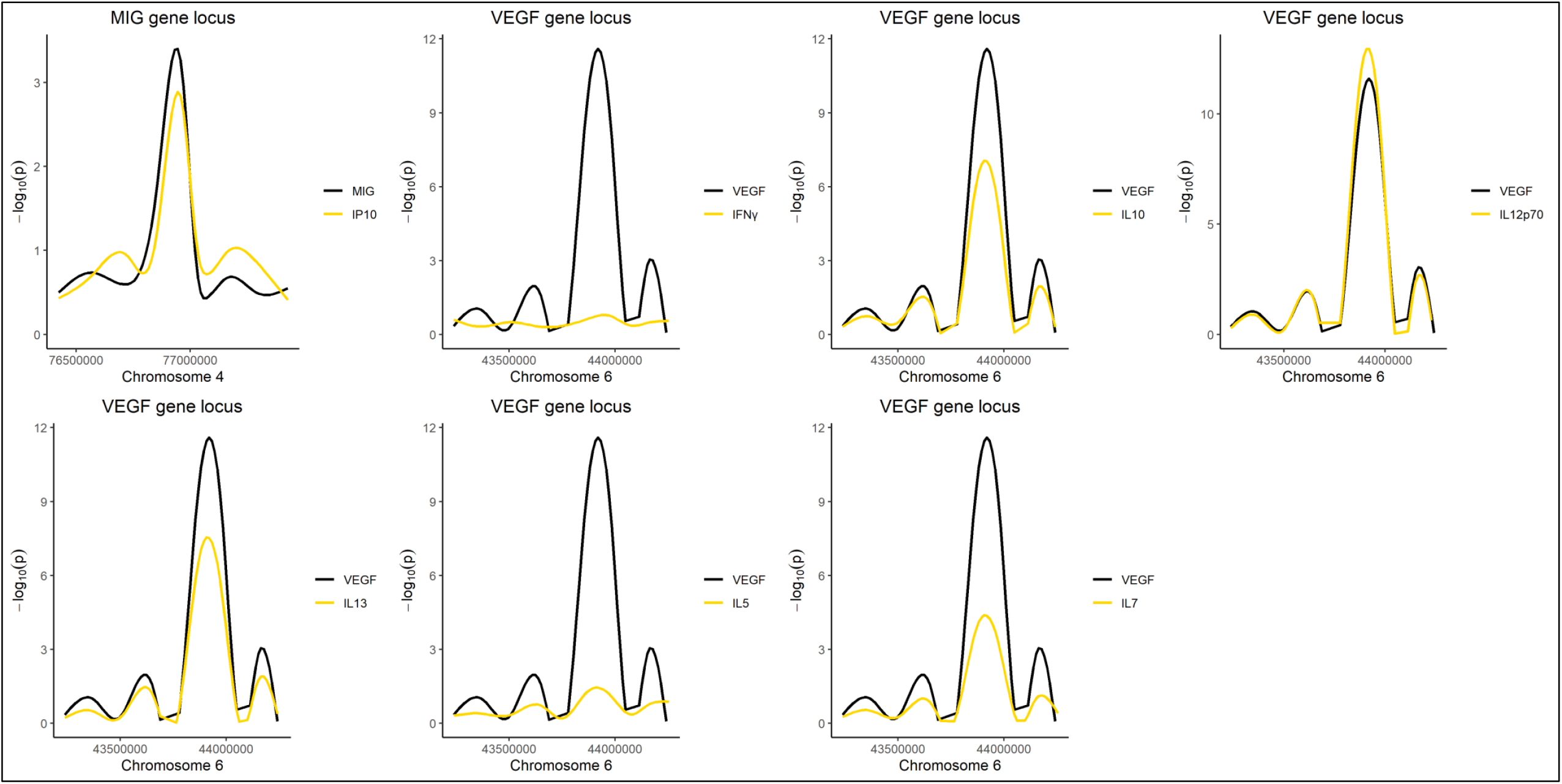
Colocalization for genome-wide association study summary statistics between selected cytokine pairs. Cytokine pairs that showed MR evidence for both causality (*P* < 0.0011 (0.05/number of cytokines)) and colocalization (posterior probability > 0.9) are plotted, within ±500 kb of the gene locus of the exposure cytokine. The y-axis of –log_10_(*p*) values is smoothed using generalized additive models.

We finally conducted MR to examine the effect of circulating cytokine levels on 20 disease traits using outcome summary statistics from largest available GWAS, including cancer, cardiovascular, metabolic and neuropsychiatric outcomes (Supplementary Table 5). All these traits are previously shown to be related to inflammation(2). Using the *cis*-pQTL instrument selection criteria and applying a Bonferroni correction for 20 outcomes, there was evidence of an association (*P* < 0.0025) between higher genetically predicted macrophage inflammatory protein-1-beta (MIP1β) levels and increased risk of stroke, between higher genetically predicted sICAM1 levels and both increased risk of major depressive disorder (MDD) and lower waist circumference, and between higher genetically predicted TRAIL levels and both increased risk of coronary artery disease (CAD) and decreased risk of breast and prostate cancer (Figure 5, Supplementary Figure 50, Supplementary Table 4). Using the *cis*-eQTL criteria, there was evidence of an association between higher genetically predicted IL1α levels and lower BMI (Figure 5). For the 15 cytokines where there were three or more instrument variants available (Table 1), weighted median and MR-PRESSO sensitivity analyses produced consistent MR estimates to the main inverse-variance weighted (IVW) analysis (Methods, Supplementary Figure 51, Supplementary Table 4). We compared the results of the two types of instrumental variables for the effects of genetically predicted cytokine levels on outcome traits and found a positive correlation between the *cis*-pQTL main MR estimates and the *cis*-eQTL main MR estimates (Pearson’s correlation coefficient *r* = 0.45, *P* = 1.2×10^−7^, Figure 6).

**Figure 5.**
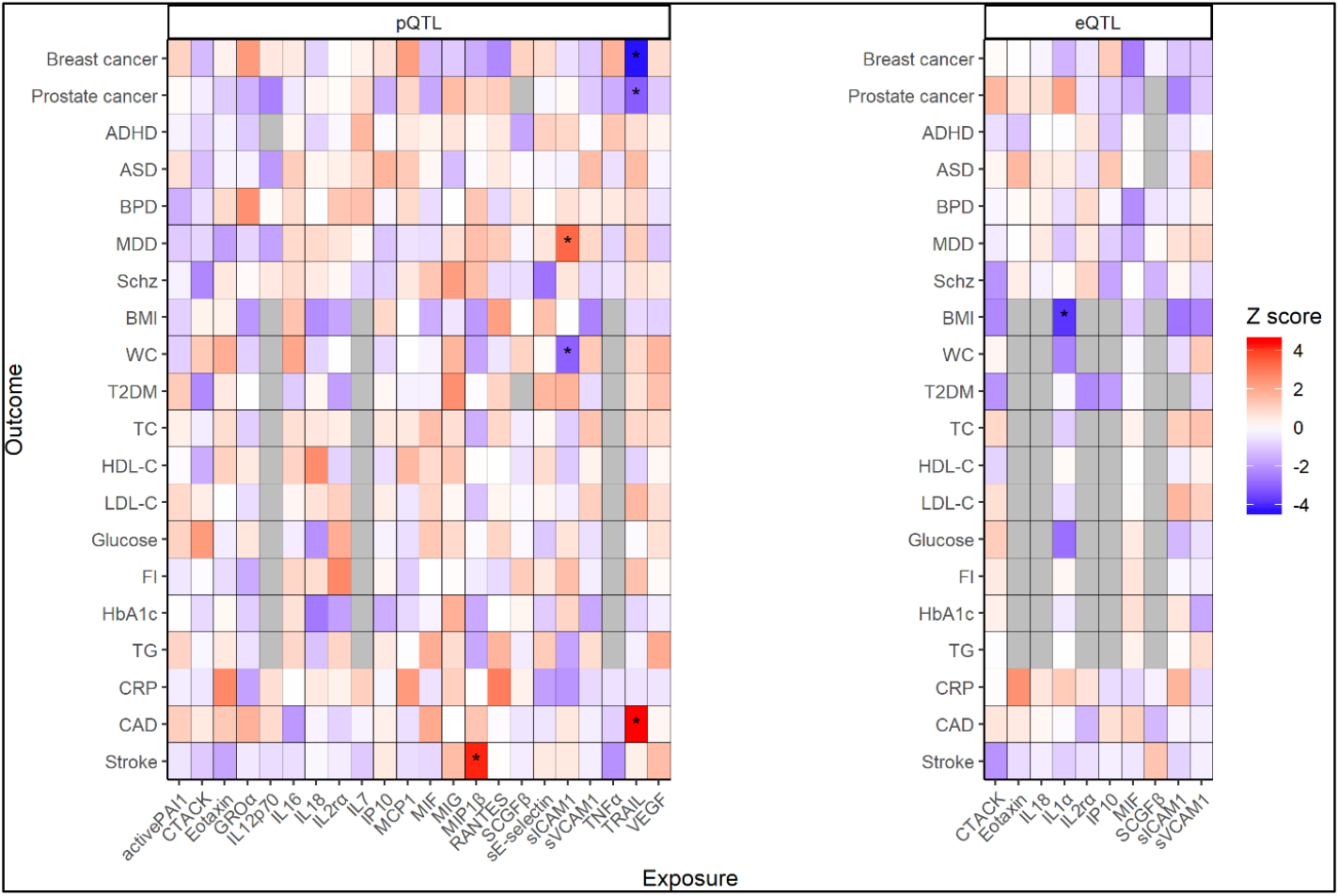
Mendelian randomization estimates for the effects of genetically predicted cytokine levels on disease outcomes when considering *cis*-pQTL (left – part A) and *cis*-eQTL (right – part B) instruments. After performing a Bonferroni correction for testing of multiple disease outcomes, associations with *P* < 0.0025 are denoted with an asterisk. pQTL: protein quantitative trait loci; eQTL: expression quantitative trait loci. ADHD: attention-deficit hyperactivity disorder; ASD: autism spectrum disorder; BPD: bipolar disease; MDD: major depressive disorder; Schz: schizophrenia; BMI: body-mass index; WC: waist circumference T2DM: type 2 diabetes mellitus; TC: total cholesterol; HDL-C: high-density lipoprotein cholesterol; LDL-C: low-density lipoprotein cholesterol; FI: fasting insulin; TG: triglycerides; CRP: C-reactive protein; CAD: coronary artery disease.

**Figure 6.**
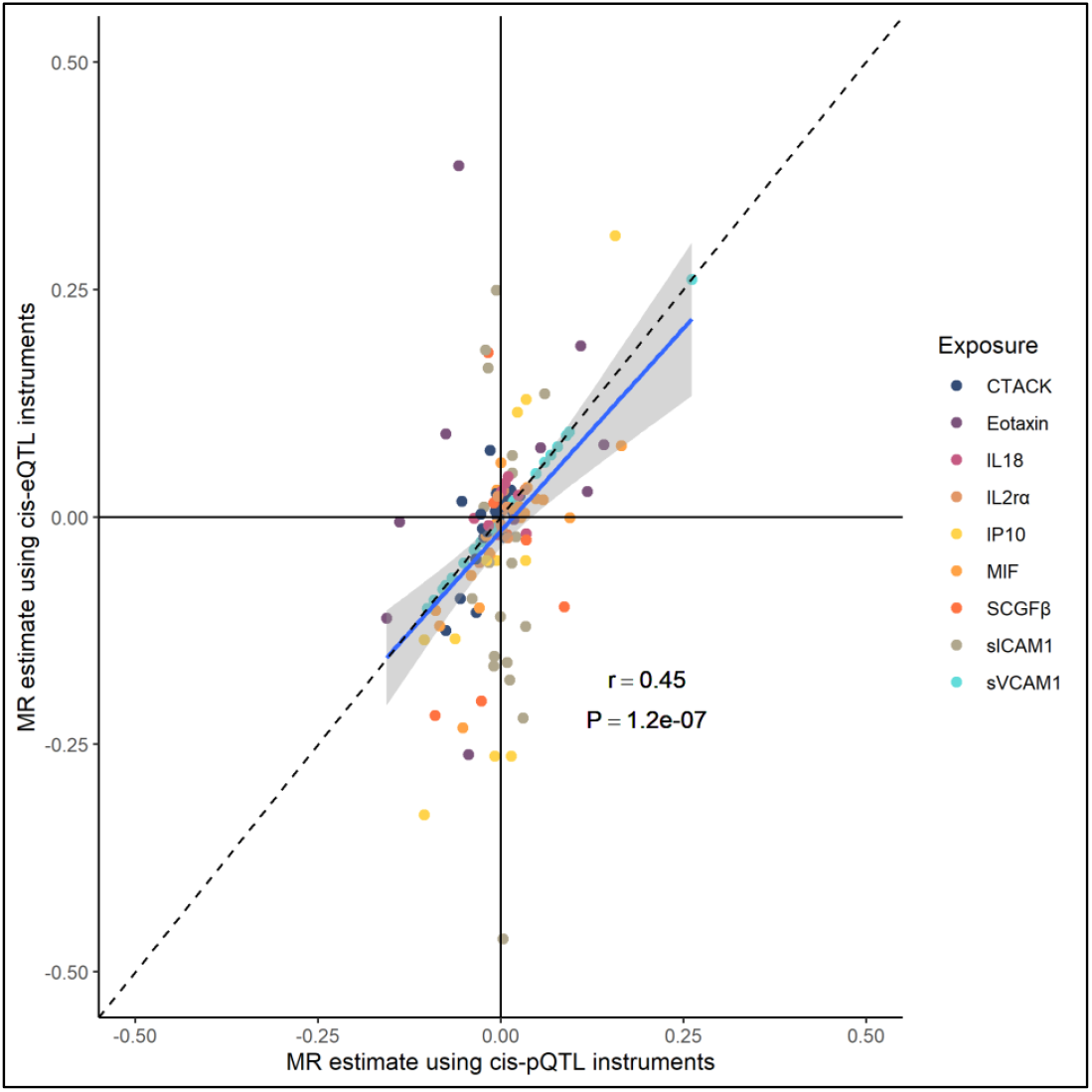
Scatter plot comparing Mendelian randomization (MR) estimates for the effects of genetically predicted cytokine levels on disease outcome traits when using the two different instrument selection criteria. The correlation between the MR estimates was quantified using Pearson’s coefficient and a linear fit between effect estimates (blue line). x-axis: MR effect estimates using *cis*-protein quantitative trait loci instrument selection; y-axis: MR effect estimates using *cis*-expression quantitative trait loci instrument selection. r: correlation coefficient; P: p value.

## Discussion

We performed a comprehensive MR analysis into the determinants, cascades, and effects of circulating cytokine levels by exploiting both pQTL and eQTL data. Our updated GWAS analyses of 47 cytokines facilitated investigation into the effects of cardiometabolic traits on systemic inflammation, and identified associations of genetically predicted BMI, SBP and smoking on circulating cytokines. GWAS of circulating cytokines in up to 13,365 healthy individuals coupled with genome-wide eQTL data of 10,361 tissue samples identified robust genetic instruments, with plausible biological relevance, for circulating cytokines. We applied these instruments in MR to explore inflammatory cascades and cytokine effects on disease risk. Our results suggest pleiotropic inflammatory effects of VEGF, and various circulating cytokines and adhesion molecules that increase risk of cancer, cardiovascular, metabolic and neuropsychiatric diseases.

We replicate associations of smoking with elevation of CRP and sICAM1, and of SBP with elevation of CRP, representing validated markers of systemic inflammation and future cardiovascular risk(16). Elevated BMI emerges as an exposure that predicts multiple indices of inflammation, including thrombosis (e.g. plasminogen activation inhibitor-1), metabolic disease and altered endothelial function or immune homeostasis (HGF, MCP1, TRAIL, sICAM1 and sE-selectin). Elevation of these mediators has been reported across human and murine studies of obesity, with fatty-acid excess and relative hypoxia emerging as drivers of the inflammatory stress response(17-19). Obesity is a leading preventable threat to global health in terms of premature cardiovascular and cancer morbidity and mortality(20), suggesting adipose tissue as a relevant setting to contextualise potential mechanisms.

To better understand cytokine regulatory networks, we examined the effects of genetically predicted circulating cytokines on levels of other cytokines, as illustrated in Figure 3. For the MR findings that could be supported by evidence of colocalization at the relevant gene locus, we provide additional support for a causal mechanism. This innovative strategy enables unravelling of inflammatory pathways and insight into mediating mechanisms, which may be of relevance where a circulating cytokine is not suitable for clinical intervention, but its upstream or downstream mediators could instead be targeted. This is illustrated by strategies targeting MCP1, highlighted in recent work as a potential target to lower large-artery stroke and cardioembolic stroke risk(21). Since the MCP1/CCL2-CCR2 axis regulates monocyte emigration from the bone marrow, prolonged therapeutic blockade may cause adverse monocyte depletion(22), prompting the need for alternative intervention strategies to overcome the depletion(23). Our results point to MCP1 affecting eotaxin expression, which would be consistent with their shared role in promotion and maintenance of the T helper cell 2 (Th2)-polarised immune response, illustrated by the co-expression of their receptors (CCR2 and CCR3, respectively) across human eosinophils(24), innate lymphoid cell, and Th2 cell subsets(25). Targeting the eotaxin/CCR3 pathway may therefore represent a novel therapeutic avenue to reduce cardiovascular risk. This precision approach appears convergent with recent mechanistic insights illuminating the contribution of eosinophils in the initiation of thrombotic events during cardiovascular disease(26).

Our approach reveals network complexity, with evidence of VEGF appearing a master regulator. This is consistent with an earlier report identifying VEGF as an upstream controller of IL12p70, IL7, IL10, and IL13^7^. We replicate and extend this analysis to reveal a wider range of cytokines within this cascade, including drivers of Th2-type responses (IL5 and IL13), TNFα, and IFNγ signalling (IFNγ, IL12) and immune resolution (IL10). With the exception of TNFα, the cytokines highlighted are notable for shared activation of the Jak-STAT signalling(27), a pathway which has evolved to sense and interpret changes in the tissue microenvironment and targetable by a range of therapeutics. Based on directionality, we infer VEGF blockade could lead to a reduction in multiple downstream molecules. Redundant angiogenic activity mediated by inflammatory and pro-angiogenic cytokines have recently been described in association with obesity, leading to treatment resistance to VEGF blockade in breast cancer(28). Thus, an overview of the cytokine regulatory network is, therefore, essential to support the discovery of common pathways and to explore cytokine redundancy. Our analysis highlights several putative cytokine networks, including the chemokines MIP1β, IL1α, and RANTES. This aligns with the shared potent pro-angiogenic effects of these molecules(29-31).

We also examined the effects of circulating cytokine levels on 20 disease outcomes. The results link circulating levels of TRAIL with CAD, and a protective effect on breast and prostate cancer risk. TRAIL was first described as a molecule capable of selectively inducing cancer cell apoptosis(32), but is now also recognised to be a potent inhibitor of VEGF-stimulated angiogenesis(33). Of therapeutic relevance, some triple-negative/basal-like breast cancer cells retain sensitivity to TRAIL as a single agent(34)whilst chemotherapeutic agents show promise in restoring sensitivity to TRAIL in hormone-refractory prostate cancer cells(35, 36). We saw evidence for associations between genetically predicted sICAM1 and increased risk of MDD, and lower waist circumference. Epidemiological studies have reported sICAM1 to be elevated in a range of neuropsychiatric disorders, but this association has been questioned due to confounders such as medication^(37)^ and smoking^(38)^. Our study links genetically predicted smoking and BMI to increased sICAM1 levels, which in turn associate with elevated risk of MDD. Conversely, our MR association between genetically predicted MIP1β levels and risk of stroke is consistent with the ability of MIP1β to induce endothelial cell adhesion, and the formation of vascular plaques associated with stroke(39).

Furthermore, using eQTL instruments, our results suggest an effect of higher IL1α levels on decreasing BMI. IL1α is active following translation(40), and some *in vitro* evidence implies IL1α attenuating differentiation of pre-adipocytes and suppressing lipid accumulation(41), suggesting biological relevance.

Thus, through analysis of genetically proxied cytokines, we suggest a pattern of pro- and anti-angiogenic and inflammatory effectors linking obesity to multi-morbidity. Given that all cytokines investigated in our study represent viable targets for pharmacological intervention^3^, these findings could have important clinical implications for the prevention of disease, and therefore warrant further investigation.

Our use of genetic instruments encompassed the cumulative lifelong effects of natural variation in circulating cytokine levels, thus making progress towards overcoming the confounding, reverse causation and measurement error biases that can cloud causal interpretation of associations identified in conventional observational research(42). This is particularly relevant given the complex interactions that underlie levels of circulating cytokines(43), and therefore similarly relates to our investigation of cytokine pathways and effects on disease risk. In our analyses, we aimed to maximise the validity of instruments selected to proxy levels of circulating cytokines by only considering variants that located at the corresponding gene locus. This strategy for selecting instruments is likely to both increase the biological relevance of the instrument, and decrease the possibility of horizontal pleiotropy(44). Our use of complementary pQTL and eQTL instrument selection criteria offered distinct but consistent findings and provided a time-efficient and cost-effective approach for identifying potential drug targets for the prevention of a broad range of common and serious diseases. Furthermore, for associations between cytokines, we used colocalization analyses to strengthen the evidence for causal effects.

There are also limitations to our work. The MR analyses should not be directly extrapolated to infer the effect of a clinical intervention^25^, as the instruments employed represent the cumulative effect of lifelong genetic predisposition, while a clinical intervention typically represents a discrete event at a particular time point(45). Our ability to assess causality between complex cytokine networks based on genetic methods remains limited by the availability of genetic instruments and the expression profiles of these molecules. Furthermore, the MR estimates may be biased by possible pleiotropic effects of the variants employed as instruments, where they influence the outcome under consideration through pathways independent of the cytokine being investigated. To minimise this risk, instruments for cytokines were selected from their corresponding gene loci to increase their specificity(44). Where possible, MR sensitivity analyses using methods that are more robust to the inclusion of pleiotropic variants were also incorporated(46), and provided consistent results. Another potential source of bias in the MR analyses relates to the use of weak instruments(47, 48). However, 29 out of 32 instruments included only variants with F-statistics > 10, the only exceptions being *cis*-eQTL instruments for IL1α, MIF and sICAM1(49). Finally, our approach considered circulating cytokine levels and cytokine gene expression aggregated across tissues, rather than restricting to biologically relevant sites, thus potentially having impact on the generalizability of our findings.

In conclusion, we used large-scale genetic data relating to cytokine levels to assess causal links between cardiometabolic traits and systemic inflammation. By incorporating gene expression data, we identified valid and biologically plausible instruments for use in MR analyses and investigated effects of circulating cytokines on inflammatory pathways and disease risk. Our findings are supported by existing work and add insight into the role of distinct inflammatory processes in the aetiology of a broad range of common cardiovascular, malignant, metabolic and neuropsychiatric diseases. Our work provides suggestive causal associations for further validation, after which efforts could be made towards identifying therapeutic opportunities in clinical practice.

## Methods

### Genome-wide association study cohorts

#### Northern Finland Birth Cohort 1966

Northern Finland Birth Cohort 1966 (NFBC1966) recruited pregnant women with expected date of delivery between 1^st^ January and 31^st^ December 1966(50). Overall, 12,055 mothers (over 96% of eligible women) were followed from pregnancy onwards, with 12,058 live-born children in the cohort. In the offspring 31-year data collection in 1997, all cohort members with known addresses in either the Northern Finland or Helsinki area were invited to a clinical examination(51). In total, data were received for 6033 participants, and DNA was successfully extracted for 5753 participants from fasted blood samples. Cytokines were quantified from overnight fasting plasma samples using Bio-Rad’s Bio-Plex 200 system (Bio-Rad Laboratories, California, USA) with Milliplex Human Chemokine/Cytokine and CVD/Cytokine kits (Cat# HCYTOMAG-60K-12 and Cat# SPR349; Millipore, St Charles, Missouri, USA) and Bio-Plex Manager Software V.4.3 as previously described(52, 53). Genotyping was conducted using Illumina HumanCNV-370DUO Analysis BeadChip (Illumina, California, USA) and imputed using Haplotype Reference Consortium imputation reference panel.

#### The Cardiovascular Risk in Young Finns Study

The Cardiovascular Risk in Young Finns (YFS) is an ongoing follow-up study of 3,596 children and adolescents aged 3, 6, 9, 12, 15, or 18 years. The subjects were randomly chosen from five university cities and their rural surroundings using Finnish population register. The baseline survey was held in 1980 and subsequent follow-up visits involving all five centres have been arranged in 1983, 1986, 1989, 2001, 2007, 2011 and 2017. The latest follow-up included also children and parents of the original participants.

Genotyping have been performed using the blood samples drawn at 2001 follow-up visit. Genotyping was performed with custom-build Illumina 670K array. The custom content replaced some poor performing SNPs on the Human610 BeadChip and added more CNV content after which the customized chip shared 562,643 SNPs with Illumina Human610 chip. Genotyping was performed for 2,556 samples. Prior to imputation, samples and probes with high missingness were excluded (MIND > 0.05 and GENO < 0.05). To exclude poorly functioning probes, we excluded SNPs deviating from Hardy-Weinberg equilibrium (HWE p-value < 1×10^−6^). To exclude related samples, we used 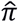 cut-off of 0.20. The pair with greater missingness was removed. After the QC steps, the data set included 2,443 individuals and 546,674 probes. The imputation was performed with IMPUTE2 software by using 1000 Genomes Phase 3 release as reference panel. After imputation, poorly imputed and rare variants (INFO < 0.7 and MAC < 3) were removed.

Biorad’s Bio-Plex Pro Human Cytokine 27-plex Assay and 21-plex Assay were used to quantify circulating concentrations of 48 cytokines from serum samples drawn at 2007 follow-up visit as previously described(54). Depending on the cytokine, imputed genotypes and cytokine concentrations were available for 116-2019 samples.

#### FINRISK

FINRISK surveys are population-based cross-sectional studies which began in 1972. A new sample is recruited every five years to monitor the health status of Finnish population. Subset of individual-level data from 1992-2012 surveys is available through THL Biobank. Cytokine quantification for FINRISK1997 and FINRISK2002 samples was performed similarly as in YFS, but quantification was done using EDTA plasma in FINRISK1997 and heparin plasma in FINRISK2002(55). In FINRISK1997, a custom 20-plex array was used in cytokine quantification. In FINRISK2002, only participants between 51 and 74 years were selected for the analysis. Imputation was performed using 1000 Genomes Phase 3 as reference panel. Poorly imputed variants (INFO < 0.7) and variants with low minor allele count (MAC < 3) were excluded. Depending on the cytokine, imputed genotypes and cytokine concentrations were available for 3440-4613 samples from FINRISK1997 and 843-1705 samples from FINRISK2002.

### Cytokine genome-wide association study

We conducted GWAS for 41 cytokines in FINRISK+YFS population and for 16 cytokines in NFBC1966 (Supplementary Table 6)(8). The data pre-processing was done in a similar manner to previous GWAS analyses(7, 8). Inverse-normal rank transformation was first applied to the traits, before regressing the transformed measures on age, sex and the first 10 genetic principal components. In contrast to the previous analyses(7, 8), we did not add BMI as a covariate, as this could potentially introduce collider bias into consequent MR analyses(56). The inverse-normal rank transformation was again applied to the residuals of this regression, and these transformed residual estimates were used as response variables in the GWAS. The GWAS was conducted in each study using an additive genetic model with SNPTEST2 software(57). The results for variants which showed poor imputation quality (model info < 0.7) or low minor allele frequency (MAF, < 0.05) were discarded. For the ten cytokines available in both NFBC1966 and FINRISK+YFS (Supplementary Table 6), the summary statistics were pooled by inverse variance weighted fixed-effects meta-analysis using Metal software(58).

Gene expression GWAS summary statistics were obtained from the GTEx project (release version 7), and related to 10,361 samples from a multi-ethnic group of 635 individuals(9). Results from 53 tissues were pooled using fixed-effects meta-analysis to produce cross-tissue estimates of association with gene expression(9).

### Metabolic trait instrument selection

The metabolic traits considered were BMI, low-density lipoprotein cholesterol (LDL-C), lifetime smoking (referred to as smoking), systolic blood pressure (SBP) and type 2 diabetes mellitus (referred to as diabetes). Genetic association estimates for BMI were obtained from the GIANT Consortium GWAS meta-analysis of 806,834 individuals of Europeanancestry(59). Genetic association estimates for fasting serum LDL-C were obtained from the Global Lipids Genetic Consortium GWAS of 188,577 individuals of European-ancestry(60). Genetic association estimates for smoking were obtained from a GWAS of 462,690 European-ancestry individuals, with a continuous measure of smoking created based on self-reported age at initiation, age at cessation and cigarettes smoked per day(61). Genetic association estimates for SBP were obtained from a GWAS of 318,417 White British individuals, with correction made for any self-reported anti-hypertensive medication use by adding 10mmHg(62). Genetic association estimates for liability to type 2 diabetes mellitus came from the DIAGRAM Consortium GWAS meta-analysis of 74,124 cases and 824,006 controls of European ancestry(63). Instruments for each trait were selected as single-nucleotide polymorphisms (SNPs) that associated with that trait at genome-wide significance (*P*<5×10^−8^) and were in pair-wise linkage disequilibrium (LD) *r*^2^<0.001. All clumping was performed using the TwoSampleMR package in R(64).

### Cytokine instrument selection

Two distinct instrument selection criteria were used to identify variants to proxy the effect of circulating cytokine levels in MR analyses. Firstly, variants located within 500 kb of the gene locus corresponding to a cytokine under study that also related to circulating levels of that cytokine with association *P*-value < 1×10^−4^ were selected. These are referred to as *cis*-pQTL. Secondly, variants located within 500 kb of the gene locus corresponding to a cytokine under study that related to both expression of that gene at *P* < 1×10^−4^ and circulating levels of that cytokine at *P* < 0.05 were selected. These are referred to as *cis*-eQTL. The gene locations were extracted per human genome build 19 (released February 2009) using UCSC Genome Browser (accessed on 18^th^ June 2019) (65). Only variants for which both exposure and outcome genetic association estimates were available for any given MR analysis were considered as potential instruments for that analysis. All variants were clumped to a pairwise linkage disequilibrium threshold of *r*^2^<0.1 using the TwoSampleMR package in R(64). The F statistic was calculated as a measure of the strength of the instruments(66).

### Outcome genetic association estimates

Disease outcomes falling into the categories of cardiovascular (CAD and stroke), cancer (breast and prostate), metabolic (BMI, fasting glucose, fasting insulin, type 2 diabetes mellitus, low-density lipoprotein cholesterol, high-density lipoprotein cholesterol and triglycerides) and neuropsychiatric (attention-deficit hyperactivity disorder, autism spectrum disorder, bipolar disorder, major depressive disorder and schizophrenia) traits were investigated, based on the availability of large-scale GWAS summary data. In addition, the effects of circulating cytokine levels on C-reactive protein levels, an established biomarker of inflammation, were investigated. Trait definitions and details of the original GWAS analyses are provided in Supplementary Table 5.

### Mendelian randomization analysis

MR analysis was performed using the two sets of instruments (*cis*-pQTL and *cis*-eQTL) to investigate effects of genetically predicted circulating cytokine levels on levels of other circulating cytokines (Supplementary Table 6) and the considered disease outcomes (Supplementary Table 5). The Pearson correlation coefficient was used to estimate the similarity between MR estimates obtained for the same exposure-disease outcome associations when using the two different instrument selection criteria (pQTL and eQTL). The ratio method was used to obtain MR estimates, with first order weights used to generate standard errors(67). Where more than one instrument variant was available for a given analysis, MR estimates obtained from different instruments were pooled using the multiplicative random-effects inverse-variance weighted (IVW) MR method. MR is prone to bias when the genetic variants used as instruments affect the outcome through some pathway that is independent of the exposure under consideration, a phenomenon termed pleiotropy(68). To explore this possibility, MR methods that make distinct assumptions on the inclusion of pleiotropic variants were performed in sensitivity analyses where three or more instrument variants were available. Specifically, the weighted median and MR-PRESSO methods were used(69, 70). Briefly, the weighted median approach orders the MR estimates produced by each instrument by their magnitude weighted for their precision and produces an overall MR estimate based on the median value, with standard error estimated by bootstrapping(69). It is a consistent approach when more than half of the weight for the analysis is derived from valid instruments (69). The MR-PRESSO detects outliers using the squared residuals from the regression of the variant-outcome association estimates on the variant-exposure association estimates with the intercept fixed to zero, and repeats such regression-based MR analysis after excluding any identified outlier variants(70). To account for multiple testing, we applied a Bonferroni correction accounting for the number of outcomes for each analysis; thus, for the MR analysis on 20 disease outcomes and 47 cytokines, the thresholds for statistical significance were 0.0025 and 0.0011, respectively. The MR sensitivity analyses were only performed to explore the robustness of the main IVW analysis to potential pleiotropy and as such no statistical sensitivity threshold was applied for these.

### Colocalization

We used GWAS-pairwise for colocalization to further evaluate the cytokine pairs for which there was MR evidence of causal effect in either direction(15). The colocalization analysis investigates whether the same variant within a genetic locus is associated with both exposure and outcome trait. We used the *Z* scores from the cytokine GWAS summary statistics. The number of variants included in each analysed chunk was set automatically by using an approximately linkage disequilibrium independent block file(15). Results with a posterior probability of association > 0.9 within the gene locus of the putative causal cytokine of each pair were deemed statistically significant. We only considered the gene region for the exposure cytokine, and thus any evidence for colocalization would imply supporting evidence for our MR results.

### Data availability and ethical approval

All supporting data for this work are available within the article, its supplementary files and the citations provided. *The GWAS summary statistics generated in this work will be made publicly available upon the publication of the article*. All study cohorts used in this work had already obtained relevant ethical approval and written participant consent. Statistical analysis was performed using R (version 3.6.0). The software code for the analyses are available from the authors upon request.

## Data Availability

All supporting data for this work are available within the article, its supplementary files and the citations provided. The GWAS summary statistics generated in this work will be made publicly available upon the publication of the article. All study cohorts used in this work had already obtained relevant ethical approval and written participant consent.

## Acknowledgements

We thank all cohort members and researchers who participated in the NFBC1966 31-year study. We also wish to acknowledge the work of the NFBC project centre. The authors acknowledge the contributors of the data used in this work: BCAC, CARDIoGRAMplusC4D, CHARGE, CKDGen, DIAGRAM, GIANT, GLGC, MEGASTROKE, PRACTICAL and UK Biobank.

